# Effect of workplace infection control practices on workers’ psychological distress: a large-scale cohort study during the COVID-19 second state of emergency in Japan

**DOI:** 10.1101/2021.10.05.21264597

**Authors:** Toyohiko Kodama, Tomohiro Ishimaru, Seiichiro Tateishi, Ayako Hino, Mayumi Tsuji, Akira Ogami, Tomohisa Nagata, Shinya Matsuda, Yoshihisa Fujino

**Author notes:** Corresponding author: Yoshihisa Fujino M.D., M.P.H., Ph.D., Department of Environmental Epidemiology, Institute of Industrial Ecological Sciences, University of Occupational and Environmental Health, Japan, 1-1, Iseigaoka, Yahatanishiku, Kitakyushu, 807-8555, Japan.

## Abstract

**Background:** The COVID-19 pandemic has dramatically transformed the work environment and work practices worldwide. Long-term infection control practices may increase the psychological stress on workers, and conversely, inadequate infection control practices at the working place may increase the fear of infection. This study aimed to determine the relationship between infection control practices at the workplace and employee mental health during the COVID-19 pandemic in Japan.

**Methods:** This study was conducted in December 2020 and February 2021. The participants had undergone a preliminary survey, which revealed that they were in good mental health. Their psychological distress was investigated via a second survey, and the factors associated with distress were studied using a logistic model.

**Results:** The results of the second survey indicated that 15.1% of participants demonstrated psychological distress. This was associated with leave-of-absence instructions, instructions for shortening business hours, and requests to avoid the workplace in case of any symptoms.

**Conclusion:** The study found that while some infection control practices reduce workers’ distress, others worsen it. Employers need to consider infection control practices as well as the worsening mental health of employees following a decrease in income caused by such measures. Follow-up studies may be necessary to clarify the long-term effects on workers’ mental health.

## Background

The COVID-19 pandemic has brought about significant changes in public health, particularly in mental health. Fear of infection, unstable employment, and economic conditions, as well as counter-measures against infection such as avoidance of physical contact and restrictions on movement have reduced opportunities for social interaction; this has a deteriorating effect on the mental health of the population. Previous studies showed increased anxiety and mental burdens in areas where lockdowns have been ordered.^1^ Other negative effects associated with lockdown include worsening of mental illnesses, depression, alcohol dependency, and suicide.^2-4^

Along with healthcare, the COVID-19 pandemic has also dramatically transformed the work environment and work practices.^5-7^ Various measures were implemented to prevent the COVID-19 infection at the working place, including mask-wearing, physical distancing, daily health checks, personal hygiene such as washing hands, and work from home. The implementation of appropriate infection control practices at the working place may positively affect the mental health of workers by creating a safe environment, which has been reported to reduce anxiety and depression.^8-9^ However, many infection control practices are efforts to maintain physical distance and reduce social contact, which have been associated with loneliness and mental distress.^10-11^ While infection control practices in general yield both positive and negative results, their effect on workers’ mental health remains to be determined.

A previous study has shown that the mental health of the Japanese deteriorated during the early stages of the COVID-19 pandemic.^12^ This study by Kikuchi et al., was a longitudinal survey of Japanese mental health from February, 2020 to April, 2020.^12^ However, the number of people infected during that period is about one-tenth of the number during the peak period, which leads to a gap in existing research. Additionally, no studies about workers’ mental health were conducted during the peak of the outbreak in Japan, which experienced a rapid spread of the infection since January 2020. For instance, the third wave of infection struck Japan in December 2021, leaving over 7,000 people infected people per day. However, as far as we are aware, there are no cohort studies that surveyed workers’ mental health after the third wave. An increase in the number of infected people would have a serious impact on employment and the economy, forcing workers to take long-term measures to prevent infectious diseases in their working places. While long-term infection control practices may increase psychological stress on workers, inadequate infection control practices at the working place may increase the fear of infection.

The aim of this study is to determine how infection control practices at the working place affect workers’ mental health during the rapid spread of COVID-19 and examine the association between infection control practices and the subsequent impact on the mental health of workers.

## Material and Methods

Ours was a prospective cohort study using an online questionnaire that focused on Japanese workers during the pandemic. The baseline survey was conducted from December 22nd to 26th, 2020 in Japan, during the beginning of the third wave of the pandemic. We have already reported details from the Protocol for our study.^13^ Research data was gathered from participants, who had employment contracts at the time of this study. The participants’ data was allocated by sex, prefecture, and occupation. We excluded inappropriate data according to the following criteria: response time of 6 minutes or less, height under 140 cm, body weight under 30 kg, inconsistent answers to similar questions throughout the survey, and wrong answers to a staged question were used to identify fraudulent responses (choose the third largest number from the following five numbers). As a result, from the initial 33,302 participants, only 27,036 were included in this study. After the baseline survey, we followed the cohort, and conducted a follow-up survey on the days, February 18–19, 2021.

This study was approved by the Ethics Committee of the University of Occupational and Environmental Health, Japan (R2-079 and R3-006).

### Assessment of worker’s mental health

To assess workers’ mental health, we used the Kessler Psychological Distress Scale (K6)^14^ at baseline and the follow-up survey. A follow-up study was conducted on February 18-19, 2021., in which the Japanese version’s validity of K6 was confirmed.^15-16^ In the current study, the cutoff for psychological distress was a K6 score of 5 or higher.

### Infection control against COVID-19 at working place

We investigated the status of infection control against COVID-19 in the participants’ working place in the follow-up study. We examined the presence of instructions from the working place regarding infection control following the re-declaration of the state of emergency in January 2021. The survey items about infection control in the working place covered leave-of-absence instructions, instructions for shortening business hours, limits to business travel, prohibitions against eating together, instructions for wearing a mask, instructions to disinfect thoroughly with alcohol when entering and leaving rooms, recommendations for daily temperature checks, encouragement of telecommuting, and requests not to come to work if not feeling well.

### Other covariates

We obtained information on participants’ profiles, characteristics, and socioeconomic status of the company they worked at, in the baseline survey. The follow-up survey items contained the following factors: sex, age, marital status, number of employees, job type, and education.

### Statistics

In the baseline survey, 7766 participants had a K6 score of 5 or higher. We excluded the 7766 participants with a K6 score of 5 or higher at baseline, since our study focused on workers who had demonstrated robust mental health in the baseline survey but then deteriorated as evidenced in the follow-up survey. After excluding inappropriate responses and workers who were unemployed by the follow-up survey and adding those who reported a healthy mental state in the baseline survey, 12,022 workers were included in the analysis. This was followed by an analysis of the changes in the mental health of the participants, which were evidenced by the follow-up survey responses (see a flow diagram of the study in Figure 1).

**Figure 1.**
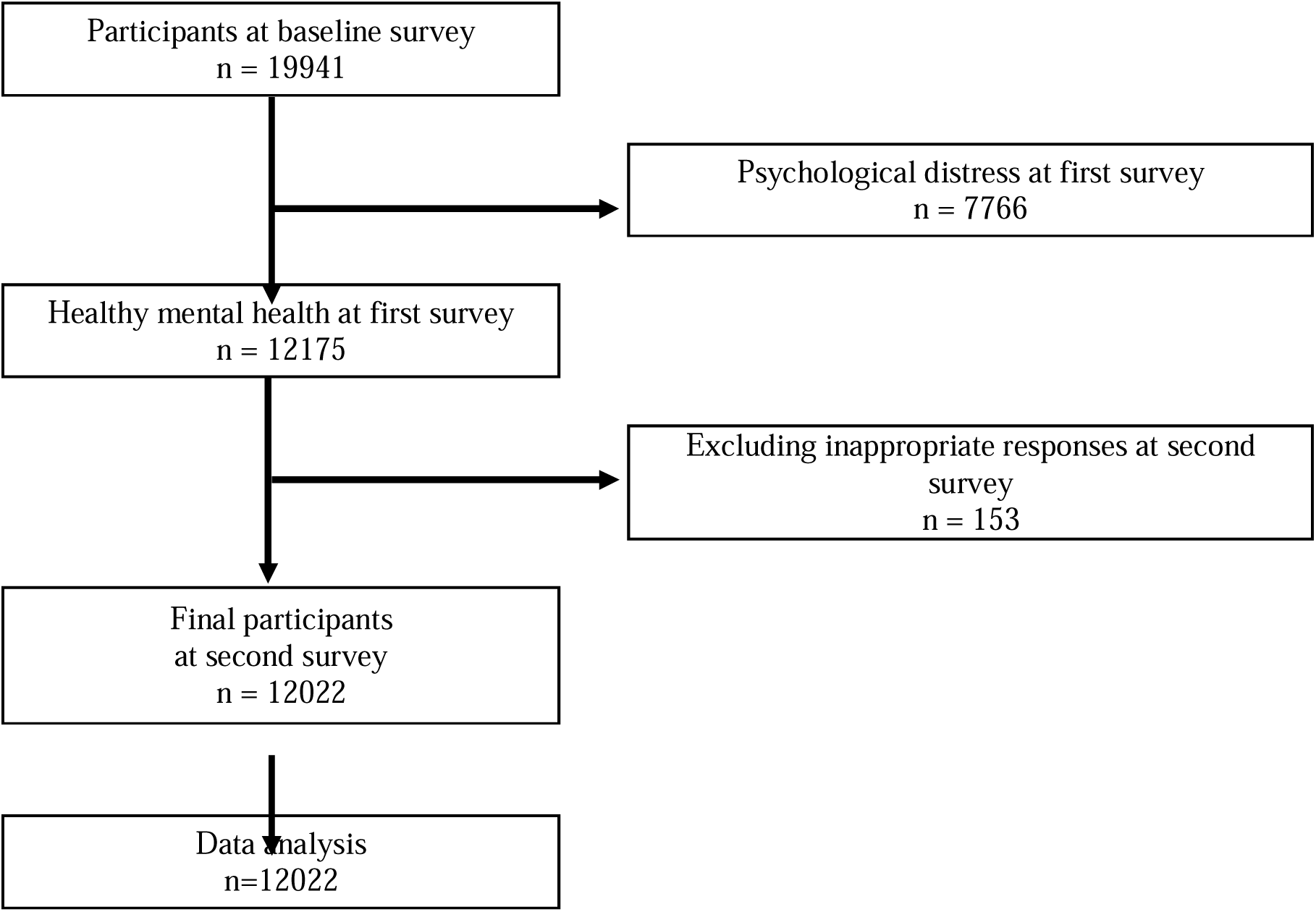
Flow diagram of the study

Odds ratios (ORs) for psychological distress and instructions from working place regarding infection control were estimated using a logistic model. Odds ratio was calculated by introducing all the instructions at the same time. Psychological distress was defined as a K6 score of 5 or higher. The multivariate model was adjusted for age, sex, marital status, number of employees, job type, and education. Moreover, we hypothesized that the ORs for workers’ distress were associated with the number of infection control practices in the working place. The number of measures were classified with reference to the analysis of the baseline survey; 0-1, 2-3, 4-5, ≧6 (more than half).^17^ Working place measures to curb infection at the baseline involved: limiting business trips, restricting the number of visitors and people at social gatherings and dinners, limiting face-to-face meetings, wearing masks during working times, installing partitions and consideration of working place layout, checking the temperature every day, encouraging employees to telecommute, encouraging them to of eat at their own desks, and requesting that employees do not come to work if they are not feeling well. A p-value of less than 0.05 was considered statistically significant. We used SPSS ver. 22 for Windows (IBM Corp., Tokyo, Japan) for analysis.

## Results

The follow-up survey found that of the 12,022 participants, 1,842 (15.1%) exhibited psychological distress. Table 1 shows the characteristics of the participants whose responses were recorded regarding the number of infection control practices (age, K6 score, sex, marital status, job type, education). Of the 12022 participants, 7373 (61.3%) indicated that they had ≥4 infection control at the working place. On the other hand, 3175 (26.4%) of the workers answered having ≤1 infection control at the working place. Participants in working places with a higher number of infection control had a higher percentage of being married and having a vocational school degree or higher.

**Table 1.** Characteristics of the participants according to the number of working place COVID-19 infection control practices

| mean(SD) or % | Number of workplace COVID-19 infection control practices<br>(n=12022) |  |  |
| --- | --- | --- | --- |
| | 0-1 | 2-3 | $\geq 4$ |
| Number of participants | 3175 | 1474 | 7373 |
| Age mean(SD) | 50.2 (9.6) | 49.7 (9.9) | 49.6 (9.86) |
| K6 mean(SD) | 2.1 (4.1) | 1.73 (3.3) | 1.95 (3.7) |
| Sex, female | 1160 (36.5%) | 646 (43.8%) | 2990 (40.6%) |
| Marital status, |  |  |  |
| Married | 1772(55.8%) | 854(57.9%) | 4707(63.8%) |
| Divorced or deceased spouse | 349(11.0%) | 167(11.3%) | 621(8.4%) |
| Unmarried | 1054(33.2%) | 453(30.7%) | 2045(27.7%) |
| Number of employees in the workplace |  |  |  |
| 1-29 | 1978(62.3%) | 729(49.5%) | 1443(19.6%) |
| 30-99 | 384(12.1%) | 256(17.4%) | 1102(14.9%) |
| 100-999 | 452(14.2%) | 279(18.9%) | 2335(31.7%) |
| $\leq 1000$ | 361(11.4%) | 210(14.2%) | 2493(33.8%) |
| Job Type |  |  |  |
| Mainly desk work | 1625(51.2%) | 675(45.8%) | 4194(56.9%) |
| Jobs mainly involving interpersonal communication | 687(21.6%) | 376(25.5%) | 1740(23.6%) |
| Mainly labor | 863(27.2%) | 423(28.7%) | 1439(19.5%) |
| Education |  |  |  |
| Junior high school | 59(1.9%) | 25(1.7%) | 52(0.7%) |
| High school | 998(31.4%) | 423(28.7%) | 1645(22.3%) |
| Vocational school/college, university, graduate school | 2118(66.7%) | 1026(69.6%) | 5676(77.0%) |

Table 2 shows the number of implemented infection control at the working place and the details. “Instructions for wearing a mask” (66.7%) was the most common infection control practices, followed by “thoroughly disinfect with alcohol when entering and leaving rooms” (64.0%). In contrast, the least common infection control practices was “instructions for leave of absence” (9.1%), followed by “instructions for shortening business hours” (10.2%).

**Table 2.** Implemented COVID-19 infection control practices in the working place

|  | Number of working place COVID-19 infection control practices (n=12022) |  |  |  |
| --- | --- | --- | --- | --- |
| | 0-1 | 2-3 | $\geq 4$ | Total |
| Number of participants | 3175 | 1474 | 7373 | 12022 |
| Instructions for leave of absence | 25 (0.8%) | 63 (4.3%) | 1002 (13.6%) | 1090 (9.1%) |
| Instructions for shortening business hours | 21 (0.7%) | 85 (5.8%) | 1122 (15.2%) | 1228 (10.2%) |
| Refrain from or limit business travel | 38 (1.2%) | 171 (11.6%) | 4828 (65.5%) | 5037 (41.9%) |
| Refrain from eating together | 55 (1.7%) | 404 (27.4%) | 6459 (87.6%) | 6918 (57.5%) |
| Instructions for wearing a mask | 104 (3.3%) | 892 (60.5%) | 7020 (95.2%) | 8016 (66.7%) |
| Thoroughly disinfect with alcohol when entering and leaving rooms. | 65 (2.0%) | 795 (53.9%) | 6838 (92.7%) | 7698 (64.0%) |
| Recommendations for daily temperature check | 59 (1.9%) | 572 (38.8%) | 6297 (85.4%) | 6928 (57.6%) |
| Encouragement of telecommuting | 49 (1.5%) | 133 (9.0%) | 2968 (40.3%) | 3150 (26.2%) |
| Request not to come to work when you are not feeling well | 96(3.0%) | 673(45.7%) | 6916 (93.8%) | 7685 (63.9%) |

Table 3 uses the logistic model to show the association between workers’ distress and instructions from the working place regarding infection control. The multivariate model included age, sex, marital status, job type, and education. Psychological distress was strongly associated with instructions for leave of absence, instructions for shortening of business hours, and requests of not coming to work if not feeling well.

**Table 3.**
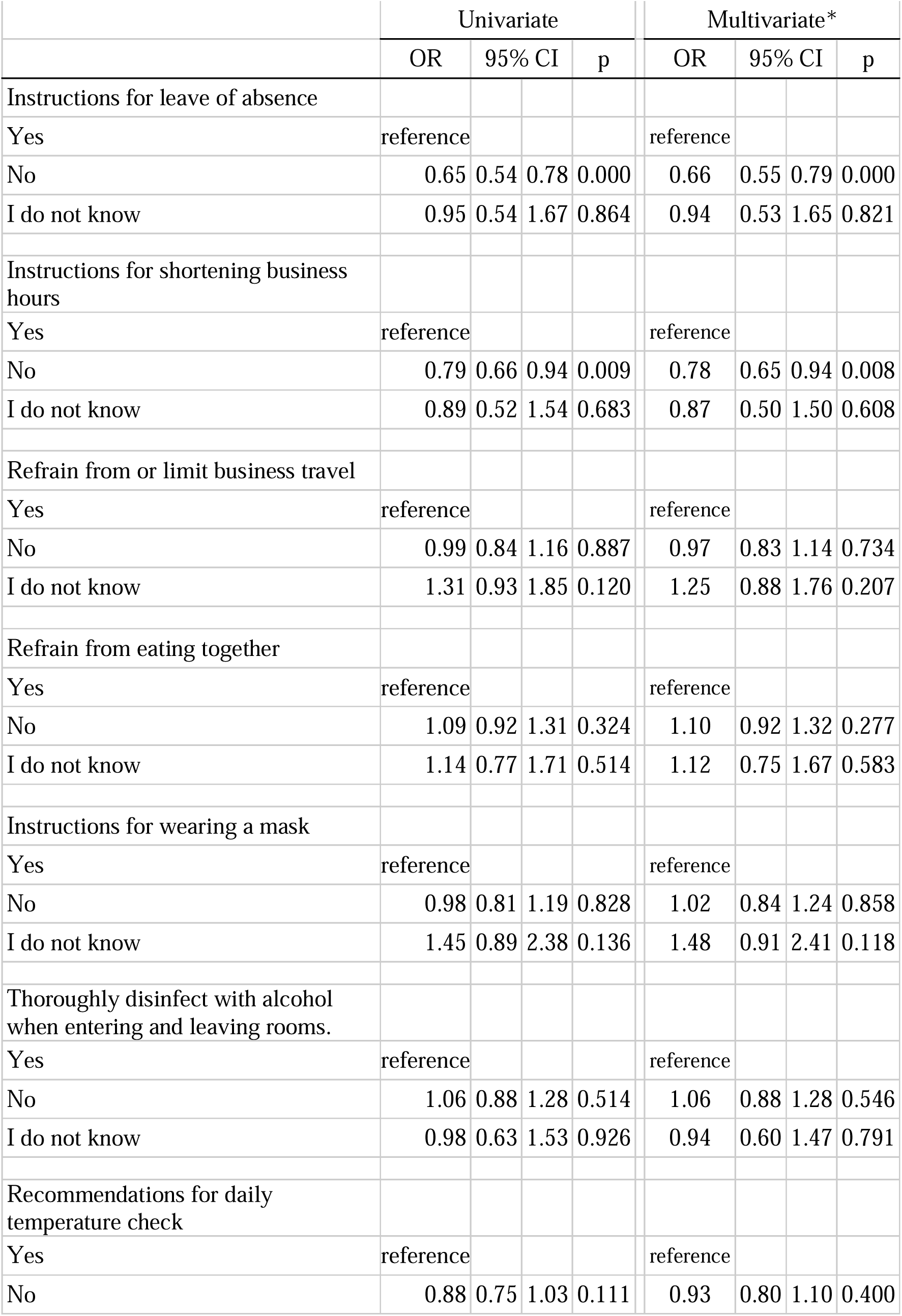
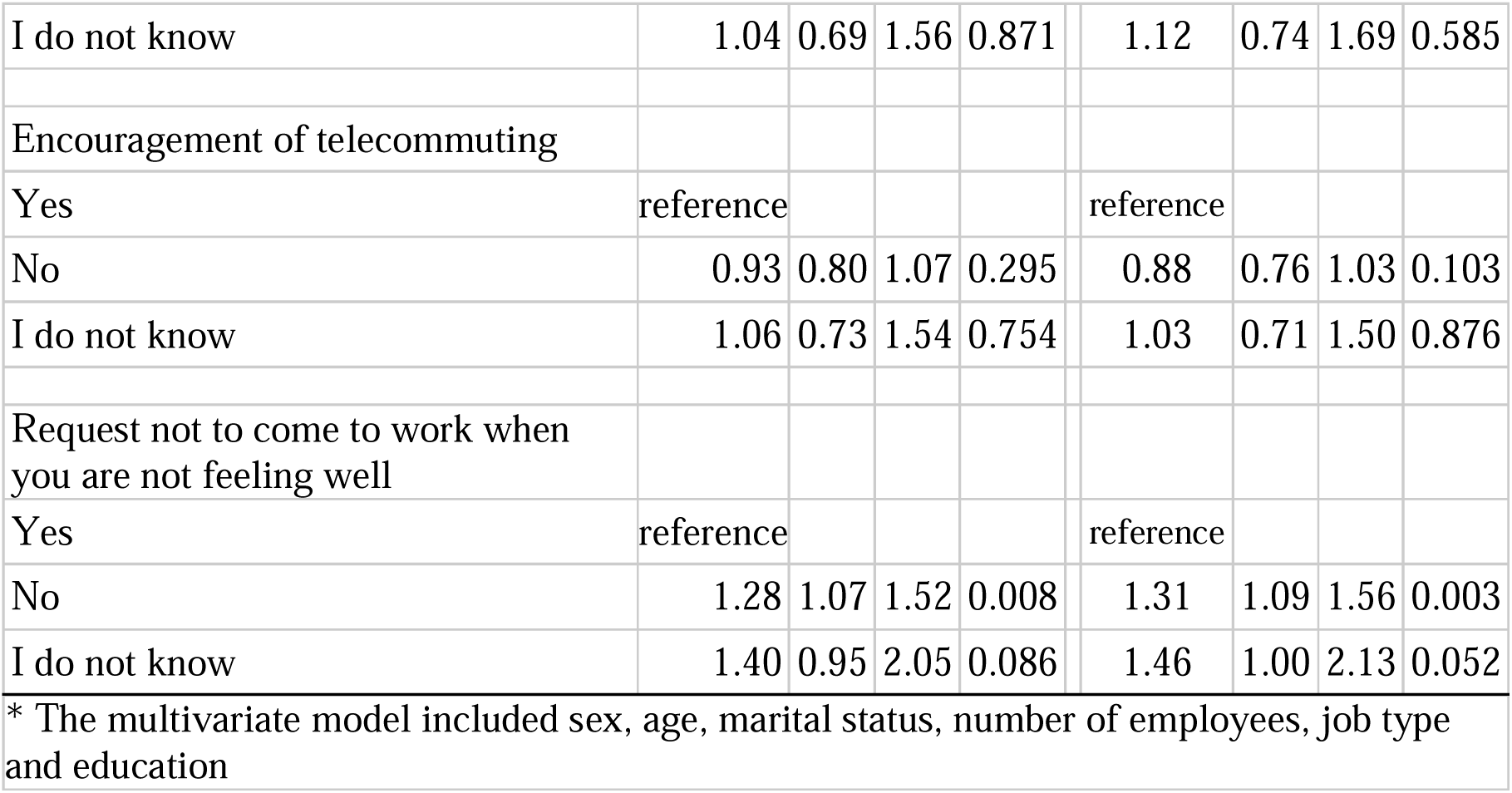
Association between psychological distress and instructions from the working place regarding infection control

Participants who answered “No” to the question about instructions for leave of absence had significantly lower ORs (OR = 0.66, 95% CI = 0.55 – 0.79, p<0.00). Participants who answered “No” to the questions about instructions for shortening the number of business hours had significantly lower ORs (OR = 0.78, 95% CI = 0.65 – 0.94, p=0.008). Participants who answered “No” to requests to not come to work if not feeling well had significantly higher ORs (OR = 1.31, 95% CI = 1.09 – 1.56, p=0.003).

We planned to estimate the ORs for workers’ distress associated with all the aforementioned infection control practices in the working place, imposed at the same time. However, instructions for leave of absence and shortening the business hours had a significantly opposite effect on workers’ mental health compared to other infection control practices. Therefore, we excluded this data and analyzed them again. As a result, the number of measures has been reduced from 9 to 7. We categorized them according to the classification in the baseline survey.^17^ The baseline divided the ten measures into four categories 0-1, 2-3, 4-5, ≧6 (more than half), and found that the more working place measures were implemented, the better the worker’s mental health. To compare these results with the baseline survey, we divided them into categories 0-1, 2-3, 4≧(more than half) in the same way as in the baseline survey

Table 4 shows the association between the number of infection control in the working place and the distress of workers. Compared to workers whose working places implemented four or more infection control practices, the OR of workers with 0 or 1 infection control practices was 1.35 (95% CI: 1.19-1.53, p=0.001).

**Table 4.**
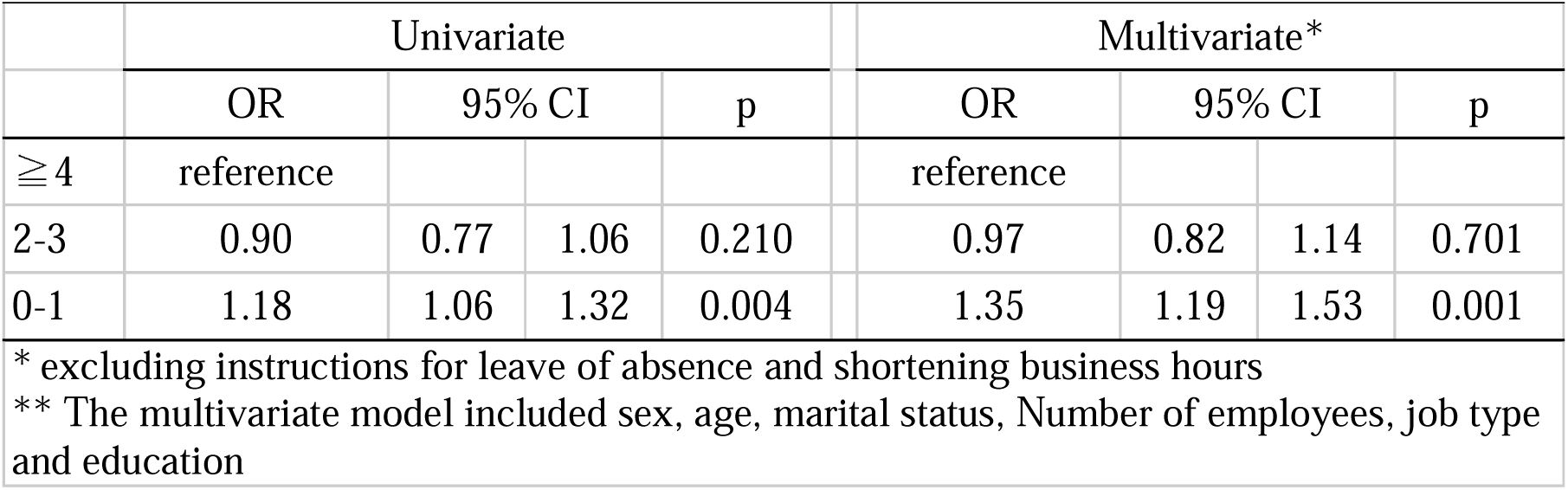
Association between psychological distress and number of workplace COVID-19 infection control practices

## Discussion

We examined the COVID-19 infection control practices in the working place during the re-declaration of the state of emergency and observed that while some control practices had a significant favorable impact on workers’ mental health, others had an unfavorable impact. In addition, workers in working place with little or no infection control practices were at a higher risk of psychological distress than workers in places with more infection control practices (other than instructions for leave of absence and shortening business hours).

This study showed that requests to “not come to work if not feeling well” were associated with a reduced risk of psychological distress. The absence of workers with poor health provides other workers a sense of security that the infection will not spread in the workplace. Such measures also allow the workers who are feeling unwell themselves to avoid the anxiety of infecting others. Sickness presenteeism is the act of going to work despite poor health, has been observed since before the COVID-19 pandemic. The reasons behind such behavior are lower income, unstable employment, guilt over increased burden on colleagues, lack of employees, and so on.^18^ Sickness presenteeism is known to be associated with poor mental health among workers.^19^ Workers who engage in frequent sickness presenteeism are reported to have a higher risk of developing depression in the future.^20^ The reasons are thought to include worsening relationship with superiors and colleagues due to decreased work efficiency and poor sleep.^20^ On the contrary, during the COVID-19 pandemic, workers will not feel conflicted about taking a leave of absence if the workplace has a clear policy of requesting to not come to work if not feeling well. In addition, the reduction of infection anxiety in the workplace will help prevent the deterioration of workers’ mental health.

Nevertheless, the instructions regarding leave of absence and shortening the business hours were associated with the worsening of workers’ distress. Perhaps the workers’ income decreased, and their economic situation worsened due to the instructions for leave of absence and shortening of business hours. Economic stress can affect mental health^21^ and low income is also associated with poorer mental health.^12, 22^ Since the leave of absence and shortening of business hours directly affects the worker’s economic situation, it may have led to increased psychological distress. The second survey of this study was conducted on the days, February 18-19th, 2021; prior to that, a state of emergency was re-declared from January 8th. In many areas, restrictions were placed on the hours of operation of restaurants, amusement centers, and other establishments that attract large numbers of people, as well as on the serving of alcoholic beverages. Since workers in these occupations are often part-timers or non-regularly employed^23^ and have lower incomes than those in regular employment,^24^ the decrease in income may have had a significant impact on psychological stress.

This study shows that, compared to working places with very few infection control practices, those with a higher number of such practices have a lower risk of causing psychological distress amongst employees. Although only one of the infection control practices was associated with workers’ mental health, taking proactive measures can have a positive impact on their mental health. There are plausible reasons for this observation. First, proactive infection control practices will reduce workers’ fears of infection from the working place, and studies have already shown that anxiety of COVID-19 infection can affect mental health.^25^ Second, the company’s proactive infection control practices will increase workers’ confidence in the workplace, leading to their psychological safety.^26^ Psychological safety is defined as individuals’ perceptions of the consequences of taking interpersonal risks in their working place,^27^ and has been shown to improve work performance, information sharing, and learning in the workplace.^28^ In addition to the above, it has also been reported to be useful in preventing the deterioration of workers’ mental health during the COVID-19 pandemic^29^ – a finding that is consistent with our view.

This study suggests that infection control practices at the working place are expected to reduce the prevalence of COVID-19 infections and are also beneficial to the workers’ mental health. In the COVID-19 pandemic, since mental health is an emergent public health issue, infection control at the working place should be encouraged as well as infection prevention and mental health support. Requests to not come to work when you are not feeling well, which have been effective for workers’ mental health, have been implemented in more than 60% of workplaces, but increased implementation is desirable. On the other hand, infection control practices that lead to a decrease in income were associated with worsening psychological distress, suggesting the need for employers to consider not only infection control practices, but also worsening mental health. It would be advisable to make careful decisions regarding instructions for leave of absence and shortening business hours, and to provide financial support as well.

However, this study has some limitations. First, due to the nature of Internet surveys, selection bias was inevitable. However, data for participants in this study was collected by a diverse selection of sex, occupation, and region to minimize participant bias. Second, because the cohort was relatively short term (3 months), it may not fully reflect the impact of infectious control practices on mental health. For example, refraining from eating together would decrease the risk of infection and reduce the fear of infection, but if the refraining is prolonged, loneliness may be exacerbated by reduced communication. Even if a measure has a positive impact on mental health at a particular time, it may have a different impact in the long term. Third, as the infection control practices are self-reported by the participants, the response may be tainted by subjective evaluation. However, we believe that misinterpretation of the answers is unlikely to occur because the options within the questions describe specific measures. Finally, the implementation status of infection control practices varies greatly depending on enterprise characteristics. Therefore, enterprise characteristics may also be an alternative indicator in terms of disease control practices. In this study, the analysis is adjusted for company size, worker occupation, and educational background. However, the possibility of the effects of unobserved enterprise characteristics cannot be excluded.

## Conclusion

This study found an association between workers’ psychological distress and infectious control practices in the workplace during the COVID-19 pandemic. Infectious control practices may have both positive and negative impacts on workers’ mental health. Requests to not come to work if not feeling well were shown to improve workers’ mental health, while infectious disease control practices that lead to reduced income were shown to worsen workers’ distress. Follow-up studies may be necessary to clarify the long-term effects on workers’ mental health.

## Data Availability

Data not available due to ethical restrictions.

## List of Abbreviations

OR: Odds Ratio
K6: Kessler Psychological Distress Scale

## Declarations

### Ethics approval and consent to participate

This study was approved by the Ethics Committee of the University of Occupational and Environmental Health, Japan (No. R2-079 and R3-006). We obtained informed consent from participants through this website.

### Consent for publication

Not applicable.

### Availability of data and material

Not applicable.

### Authors’ contributions

Y.F. was the chairperson of the study group. T.I. conceived the research questions. This research protocol was designed by all the authors and all the authors developed the questionnaire. T.K. analyzed the data with Y.F. T.K. drafted the initial manuscript. All of the authors read the initial manuscript, revised it, and approved the final manuscript.

## Acknowledgments

The current members of the CORoNaWork Project, in alphabetical order (by surname), are as follows: Professor Yoshihisa Fujino (present chairperson of the study group), Dr. Hajime Ando, Professor Hisashi Eguchi, Dr. Arisa Harada, Dr. Ayako Hino, Dr. Kazunori Ikegami, Dr. Tomohiro Ishimaru, Dr. Kyoko Kitagawa, Dr. Kosuke Mafune, Professor Shinya Matsuda, Dr. Ryutaro Matsugaki, Professor Koji Mori, Dr. Keiji Muramatsu, Dr. Masako Nagata, Dr. Tomohisa Nagata, Ms. Ning Liu, Professor Akira Ogami, Dr. Rie Tanaka, Dr. Seiishiro Tateishi, Dr. Kei Tokutsu and Professor Mayumi Tsuji. All members are affiliated with the University of Occupational and Environmental Health, Japan.

## References

1. Hyland P, Shevlin M, McBride O, Murphy J, Karatzias T, Bentall RP, Martinez F, Vallieres F. Anxiety and depression in the Republic of Ireland during the COVID-19 pandemic. Acta Psychiatr Scand. 2020;142(3):249–256. https://doi.org/10.1111/acps.13219.

2. Shi L, Lu Z-A, Que J-Y, et al. Prevalence of and risk factors associated with mental health symptoms among the general population in China during the Coronavirus Disease 2019 pandemic. JAMA Netw. Open. 2020;3(7):e2014053. doi: 10.1001/jamanetworkopen.2020.14053.

3. Yao H, Chen J-H, Xu Y-F. Patients with mental health disorders in the COVID-19 epidemic. Lancet Psychiatry. 2020;7(4):e21. https://doi.org/10.1016/S2215-0366(20)30090-0.

4. Niedzwiedz CL, Green MJ, Benzeval M, et al. Mental health and health behaviours before and during the initial phase of the COVID-19 lockdown: longitudinal analyses of the UK Household Longitudinal Study. J Epidemiol Community Health. 2021;75(3):224–231. http://dx.doi.org/10.1136/jech-2020-215060.

5. Gursoy, D, Chi, CG. Effects of COVID-19 pandemic on hospitality industry: review of the current situations and a research agenda. J. Hosp. Mark. Manag. 2020;29(5), 527-529. https://doi.org/10.1080/19368623.2020.1788231.

6. Alon, TM, Doepke, M, Olmstead-Rumsey, J, Tertilt, M. The impact of COVID-19 on gender equality (working paper). National Bureau of Economic Research. 2020. http://www.nber.org/papers/w26947.

7. Dwivedi, YK, Hughes, DL, Coombs, C, et al. Impact of COVID-19 pandemic on information management research and practice: transforming education, work and life. Int J Inf Manage. 2020;55:102211. https://doi.org/10.1016/j.ijinfomgt.2020.102211.

8. Havaei F, Ma A, Staempfli S, MacPhee M. Nurses’ workplace conditions impacting their mental health during COVID-19: a cross-sectional survey study. Healthcare (Basel). 2021;9(1):84. doi: 10.3390/healthcare9010084.

9. Cho M, Kim O, Pang Y, et al. Factors affecting frontline Korean nurses’ mental health during the COVID-19 pandemic. Int Nurs Rev. 2021 Jun;68(2):256–265. doi: 10.1111/inr.12679.

10. Benke C, Autenrieth LK, Asselmann E. Lockdown, quarantine measures, and social distancing: associations with depression, anxiety and distress at the beginning of the COVID-19 pandemic among adults from Germany. Psychiatry Res. 2020;293:113462. doi: 10.1016/j.psychres.2020.113462.

11. Giorgi G, Lecca LI, Alessio F, et al. COVID-19-related mental health effects in the workplace: a narrative review. Int J Environ Res Public Health. 2020;17(21). doi:10.3390/ijerph17217857.

12. Kikuchi H, Machida M, Nakamura I, et al. Changes in psychological distress during the COVID-19 pandemic in Japan: a longitudinal study, J Epidemiol. 2020;30(11):522–528. doi: 10.2188/jea.JE20200271.

13. Fujino Y, Ishimaru T, Eguchi H, et al. Protocol for a nationwide Internet-based health survey in workers during the COVID-19 pandemic in 2020. medRxiv. 2021:2021.02.02.21249309. https://doi.org/10.1101/2021.02.02.21249309.

14. Kessler RC, Andrews G, Colpe LJ, et al. Short screening scales to monitor population prevalences and trends in non-specific psychological distress. Psychol Med. 2002;32(6):959–976.

15. Furukawa TA, Kawakami N, Saitoh M, et al. The performance of the Japanese version of the K6 and K10 in the World Mental Health Survey Japan. Int J Methods Psychiatr Res. 2008;17(3):152–158. https://doi.org/10.1002/mpr.257.

16. Sakurai K, Nishi A, Kondo K, Yanagida K, Kawakami N. Screening performance of K6/K10 and other screening instruments for mood and anxiety disorders in Japan. Psychiatry Clin Neurosci. 2011;65(5):434–441. doi: 10.1111/j.1440-1819.2011.02236.x.

17. Yasuda Y, Ishimaru T, Nagata T, et al. A cross-sectional study of infection control measures against COVID-19 and psychological distress among Japanese workers. J Occup Health. 2021;63:e12259.

18. Takashi M. Presenteeism: research history and future tasks. Occup. Health Rev. 2020;33(1):25.

19. Rosander M. Mental health problems as a risk factor for workplace bullying: the protective effect of a well-functioning organization, Ann Work Expo Health. 2021 Jun 19;wxab040. https://doi.org/10.1093/annweh/wxab040.

20. Conway PM, Hogh A, Rugulies R, Hansen AM. Is sickness presenteeism a risk factor for depressionã A Danish 2-year follow-up study. J Occup Environ Med. 2014;56:595–603. doi: 10.1097/JOM.0000000000000177.

21. Dawel, A, Shou, Y, Smithson, M. et al. The effect of COVID-19 on mental health and wellbeing in a representative sample of Australian adults. Front Psychiatry. 2020;11(1026). doi: 10.3389/fpsyt.2020.579985.

22. Yoshioka T, Okubo R, Tabuchi T, Odani S, Shinozaki T, Tsugawa Y. Factors associated with serious psychological distress during the COVID-19 pandemic in Japan: a nationwide cross-sectional internet-based study. BMJ Open. 2021;11(7):e051115. http://dx.doi.org/10.1136/bmjopen-2021-051115.

23. Ministry of Internal Affairs and Communications. 2015 National Census. 2017.

24. Ministry of Health, Labour and Welfare. 2020 Basic Survey on Wage Structure 2021.

25. OECD. Fit Mind、Fit Job:From Evidence to Practice in Mental Health and Work、Mental Health and Work. Paris: OECD Publishing, Paris; 2015. https://dx.doi.org/10.1787/9789264228283-en.

26. Dollard MF, Tuckey MR, Dormann C. Psychosocial safety climate moderates the job demand-resource interaction in predicting workgroup distress. Accid Anal Prev. 2012;45:694–704. https://doi.org/10.1016/j.aap.2011.09.042.

27. Erkutlu H, Chafra J. Leader psychopathy and organizational deviance: the mediating role of psychological safety and the moderating role of moral disengagement. Int J of Workplace Health Manag. 2019;12(4):197–213. https://doi.org/10.1108/IJWHM-12-2018-0154.

28. Frazier ML, Fainshmidt S, Klinger RL, Pezeshkan A, Vracheva, V. (2017). Psychological safety: a meta-analytic review and extension. Personnel Psychology, 2017;70:113–165. doi: http://dx.doi.org/10.1111/peps.1218.

29. Ma Y, Faraz NA, Ahmed F, et al. Curbing nurses’ burnout during COVID-19: the roles of servant leadership and psychological safety. J Nurs Manag. 2021 Jul 14. https://doi.org/10.1111/jonm.13414.

